# Combined pre-dialysis systolic blood pressure and pulse rate assessment for all-cause and cardiovascular mortalities: A nationwide cohort study on patients undergoing haemodialysis

**DOI:** 10.1101/2024.06.30.24309750

**Authors:** Nobuhiko Joki, Tatsunori Toida, Kakuya Niihata, Ryohei Inanaga, Kenji Nakata, Masanori Abe, Norio Hanafusa, Noriaki Kurita

## Abstract

**Background and Aims:** The prognostic utility of the combined assessment of pre-haemodialysis systolic blood pressure (SBP) and pulse rate (PR) compared with their individual assessment is unclear. This study aimed to determine whether the combined assessment could enhance the prognostic utility in patients on haemodialysis.

**Methods:** This nationwide cohort study involved patients undergoing maintenance haemodialysis using the Japanese Renal Data Registry database. Exposure was defined as a combination of SBP and PR. Forty-eight levels of exposure groups were created: SBP (six levels; <100, 100-<120, 120-<140, 140-<160 [reference], 160-<180, and ≥180 mmHg) and PR (eight levels; <50, 50-<60, 60-<70 [reference], 70-<80, 80-<90, 90-<100, 100-<110, and ≥110 per minute). The primary and secondary outcomes were one-year all-cause and cardiovascular mortalities, respectively. Multivariate Cox proportional hazards models were used, and multiplicative interactions were assessed to determine the superiority of the combined model over the individual models. Additive interactions were assessed using relative excess risk due to interactions (RERI).

**Results:** The combined model explained mortality and cardiac mortality better than the individual SBP and PR models (*P*<0.001 and *P*<0.002, respectively). A lower SBP was associated with a higher risk of all-cause mortality regardless of the PR. Most categories of lower SBP or higher PR vs. the 120-<140 mmHg and 70-<80/min category had positive RERIs. Similar findings were also observed for cardiac mortality.

**Conclusions:** The combined assessment of pre-dialysis SBP and PR may help in the simple stratification of patients with excess risks that cannot be identified by individual SBP or PR assessment.

## Introduction

Pre-haemodialysis (HD) blood pressure (BP) and pulse rate (PR) are two basic physiological parameters routinely measured not only for safe and effective treatment, but also for the prediction of potential underlying cardiovascular complications and poor prognosis. A pre-HD systolic BP (SBP) of 130-160 mmHg is associated with a decrease in all-cause and cardiovascular mortalities,^1–4^ and deviation of SBP from this range exhibits a U-shaped increase in the risks characteristic of HD. Few studies examining the predictive utility of pre-HD PR for mortality suggested a dose-response increase in one-year mortality risk associated with a higher PR (>60-70 / min).^5–6^ Given that simultaneous changes in SBP and PR occur along with changes in myocardial oxygen demand and regulation of the autonomic nervous system, a combined assessment of SBP and PR could yield more accurate prognostic information. Nevertheless, very few such studies have been conducted in populations with chronic kidney disease (CKD).

A study on patients with non-dialysis CKD and hypertension demonstrated that a higher rate of pressure product (RPP), defined as the product of SBP and PR, was associated with an increased risk of major cardiovascular and cerebrovascular events and mortality, even after simultaneous adjustments for SBP and PR.^7^ The associations between high RPP and increased long-term cardiovascular and all-cause mortalities have also been reported in the general population and in patients with coronary artery disease.^8,9^ Given that RPP indicates myocardial workload and thus myocardial oxygen demand,^7^ a rationale for examining the impact of high RPP on cardiovascular clinical outcomes can be readily concurred.^10^ However, as the authors point out,^7^ if SBP and PR change in opposite directions under physiological conditions, the RPP will remain parallel,^11^ and thus this may mask the possible adverse effects associated with alterations in the respective indices. Indeed, the predictability of short-term cardiovascular disease or mortality risk due to a high shock index (SI), calculated by dividing PR by SBP, has been shown in patients with coronary artery disease and myocardial infarction, independent of SBP and PR.^12,13^ Thus, although different methods for evaluating the combination of SBP and PR are used in cases of high SBP and high PR (i.e. high RPP) and low SBP and high PR (i.e. high SI), few studies have considered both cases simultaneously. Patients undergoing maintenance dialysis who often have left ventricular dysfunction or coronary artery disease are likely to have poor clinical outcomes.

Thus, to resolve this dilemma, we examined the combined effect of SBP and PR on all-cause and cardiovascular mortalities in patients on HD using data from the Japanese Renal Data Registry (JRDR), which encompasses nationwide dialysis centres in Japan. This epidemiologic approach allows quantification of the excess risk of the combined category of SBP and PR over the individual SBP and PR indices.^14,15^

## Methods

### Study design and setting

This nationwide cohort study used data from the JRDR database established by the Japanese Society for Dialysis Therapy (JSDT). The JRDR has accumulated data of patients on dialysis retrieved through annual surveys of all dialysis units in Japan conducted at the end of each year. The details of the JRDR have been published previously.^16^ We used the JRDR data for 2019 as the baseline data and those for 2020 as the follow-up data. The response rates in 2019 and 2020 were 98.3% and 98.8% on a facility basis, and 94.5% and 95.1% on a patient basis, respectively.^17,18^

### Participants

Patients on haemodialysis (including haemodiafiltration [HDF] and haemofiltration) as of the end of 2019, aged ≥20 years were included in the study. The exclusion criteria were as follows: (1) missing SBP and PR data, (2) implausible data considered to be outliers or apparent errors (e.g. SBP <70 or >260 mmHg, diastolic blood pressure <30 or >150 mmHg, pulse pressure <15 or ≥150 mmHg, PR <30 or >180 per minute, height <120 or ≥200 cm, body weight <20 or ≥200 kg, body weight <25 kg, body mass index [BMI] <10 or >50 kg/m^2^, ultrafiltration [possibility of infusion during dialysis] <-1.0 kg, haemoglobin <6.0 g/dL, and C-reactive protein level >50 mg/dL), and (3) withdrawal from dialysis during follow-up.

### Exposures

Exposure was defined as a combination of pre-HD SBP and PR, both of which are components of the RPP or SI. In this study, we decided to examine the effects of combinations of SBP and PR categories, because we expected the effects of high SBP and low PR to differ from those of low SBP and high PR, even if they had the same RPP. SBP was categorized into the following six levels: <100, 100-<120, 120-<140, 140-<160, 160-<180, and ≥180 mmHg. PR was categorized into the following eight levels: <50, 50-<60, 60-<70, 70-<80, 80-<90, 90-<100, 100-<110, and ≥110 per minute. A total of 48 levels were created based on the 6 × 8 categories. For this study, 140-<160 mmHg and 60-<70 /min was used as the reference category.

### Outcomes

The primary outcome was all-cause mortality during the one-year follow-up period, and the secondary outcome was mortality due to heart disease during the same period. Heart diseases included heart failure, pulmonary oedema, ischaemic heart disease (IHD), arrhythmia, valvular heart disease, other heart diseases, hyperkalaemia, and sudden death. The causes of death were diagnosed and reported by the on-site physicians based on the patients’ medical records. The follow-up period started on 31 December 2019 and ended on events involving death or kidney transplantation, or on 31 December 2020 whichever occurred first. We collected data on the month of death and causes of death from the JRDR database in 2020. Because the JRDR only records the month of death, withdrawal of dialysis, and kidney transplantation without specific days, we assumed that each event occurred on the 15th day of the month for our estimations.^19,20^

### Covariates

Covariates were defined as variables considered to be determinants of SBP, PR, and mortality, and were identified based on evidence from the literature and clinical expertise through discussion among the investigators (NJ, KN, TT, KN, and NK). We selected minimally sufficient covariates to estimate the total effects of exposure using the established analysis rule with the DAGitty web application (www.dagitty.net),^21,22^ based on the directed acyclic graph model (Supplementary Figure 1). The following variables were selected as covariates: age; sex; diastolic blood pressure; interdialytic weight gain (IDWG) calculated as pre-dialysis body weight minus post-dialysis body weight; BMI; serum C-reactive protein levels; haemoglobin levels; atrial fibrillation (AF); use of antihypertensive drugs; smoking; diabetes; and history of IHD, cerebral haemorrhage, and cerebral infarction.

### Statistical analysis

For descriptive statistics, continuous variables are summarised as medians (interquartile ranges), and categorical variables are summarised as frequencies and percentages. To examine the impact of exposure on the outcomes, we used Cox proportional hazards models that included the aforementioned covariates. Our interest was whether the combination of SBP and PR could explain the impact on outcomes better than the individual SBP and PR categories. This is mathematically equivalent to examining whether the addition of 35 interaction terms from the (6-1)×(8-1) categories to the respective categories of SBP and PR is statistically significant (i.e. evaluating multiplicative interactions). Therefore, we created two separate models, one including SBP and PR categories and interaction terms and the other including SBP and PR categories, and compared the models with and without interaction terms using the Wald test.^23^ This analysis was performed for both primary and secondary outcomes. Furthermore, to evaluate the potential additive interaction between SBP and PR, we calculated the relative excess risk due to interaction (RERI) in the models, including exposures and covariates.^14,15,24^ For semiquantitative visualisation of the relationships between the three factors of pre-HD SBP and PR and the outcomes, heatmaps were overlaid on tables for the hazard ratios (HRs) and RERIs by the SBP and PR combinations.^25^ The sequential scheme “YlOrRed” colour palette was used for the HRs heatmaps, and the divergent scheme “RdBu” colour palette was used for the RERIs heatmaps.^26^ Missing values were imputed using a multiple imputation method, creating 20 datasets using conditional chained equations, and combining them according to Rubin’s rule.^27^ All analyses were performed using Stata/SE version 18 software (Stata LP, College Station, TX, USA).

### Ethics statement

The data used in this study do not contain any personally identifiable information. The present study was conducted following Japan’s privacy protection laws and ethical guidelines for epidemiological studies published by the Ministry of Education, Science, and Culture, Ministry of Health, Labor, and Welfare, and the STROBE guidelines. The study protocol was approved by the Medicine Ethics Committee of the JSDT (No.60) and conducted in accordance with the Declaration of Helsinki. The requirement for written informed consent was waived due to the retrospective nature of this study.

## Results

### Patient characteristics

A total of 321,077 patients, aged ≥20 years undergoing HD or HDF were identified. Of these, 45,418 patients with missing or implausible BP or PR values were excluded and 275,659 patients were enrolled at the end of 2019. Furthermore, 444 patients were excluded owing to missing follow-up data at the end of 2020. Finally, 275,215 patients were included in the analysis.

The patient characteristics are presented in Tables 1 and 2. In particular, patients in the SBP <100 mmHg category had a higher median age and CRP level, longer dialysis duration, lower median BMI, and an increased incidence of AF and IHD (Table 1). However, the PR ≥100 / min category had lower median age, higher median CRP, and an increased incidence of diabetes, AF, and IHD (Table 2).

**Table 1.**
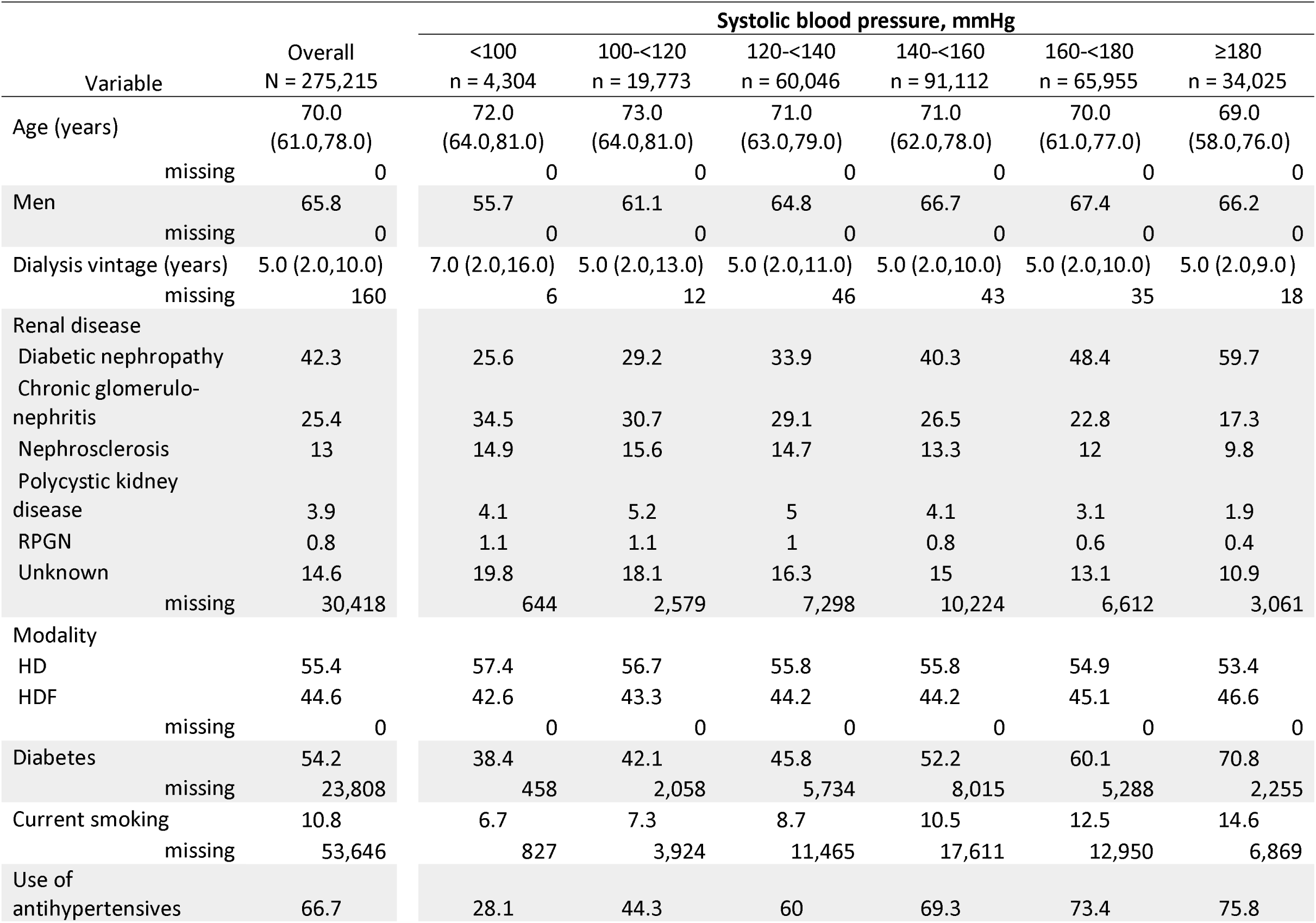
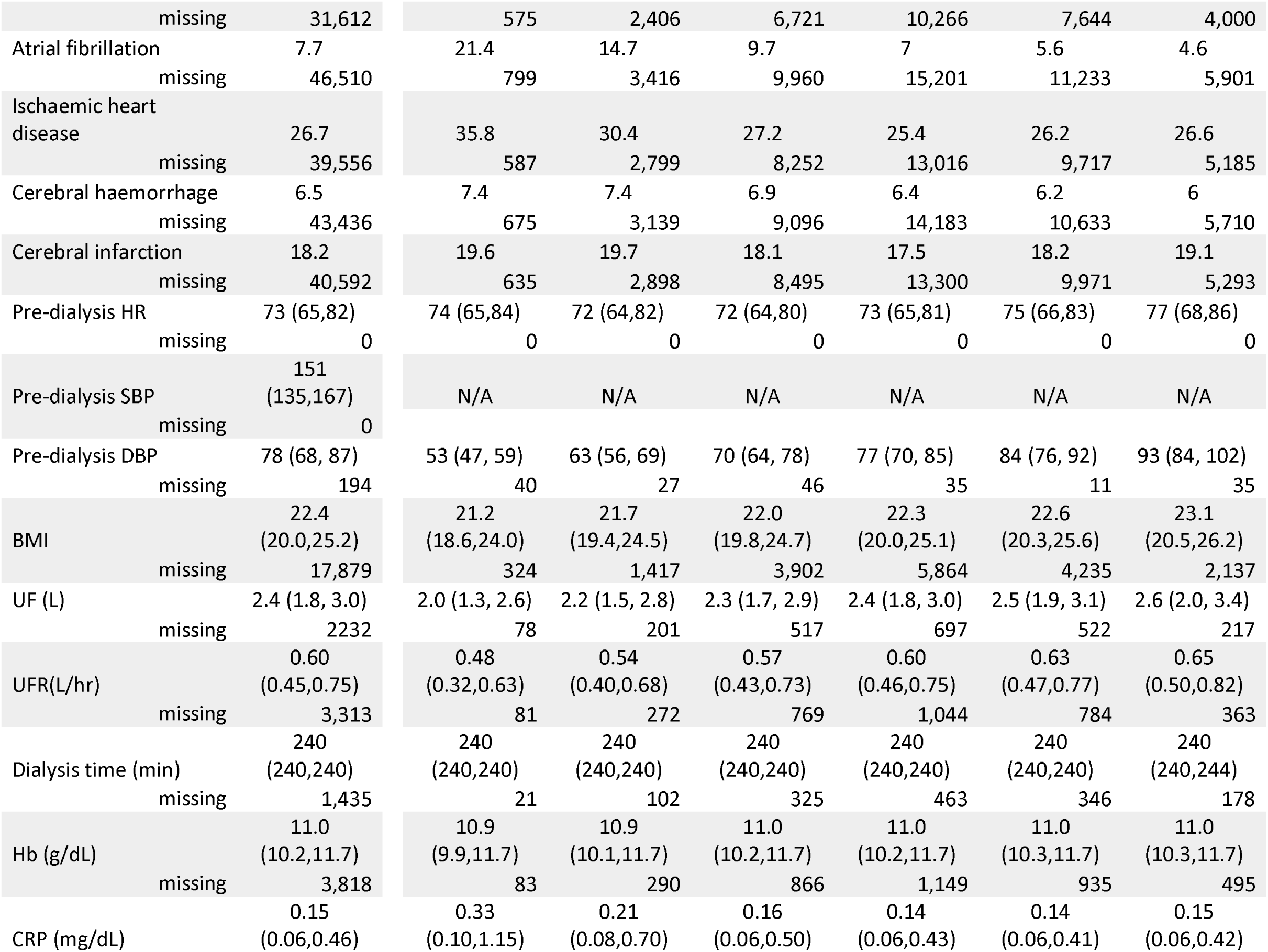
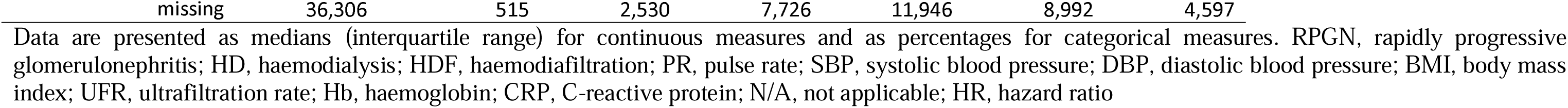
Baseline characteristics of the overall patients and those categorised by the systolic blood pressure.

**Table 2.**
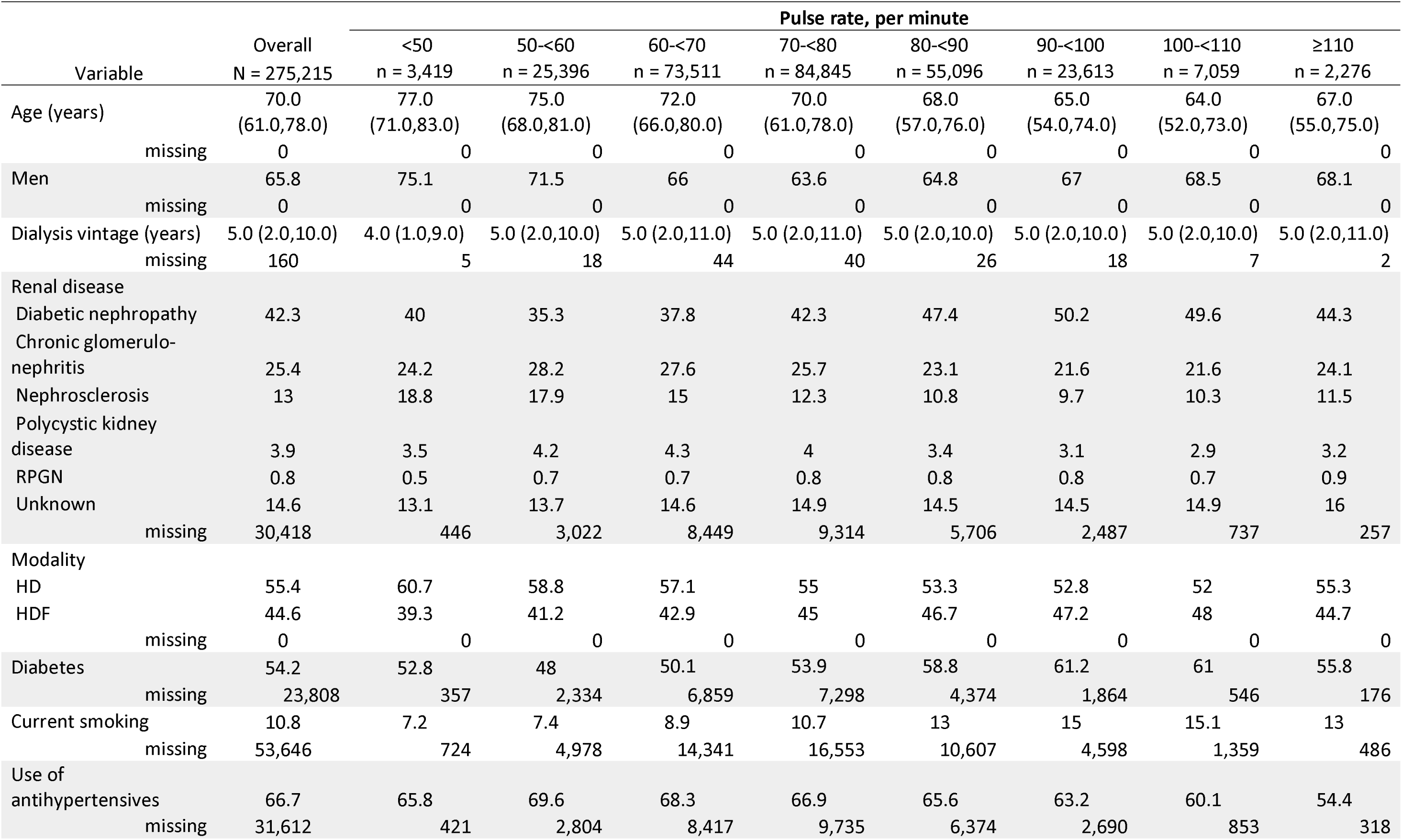
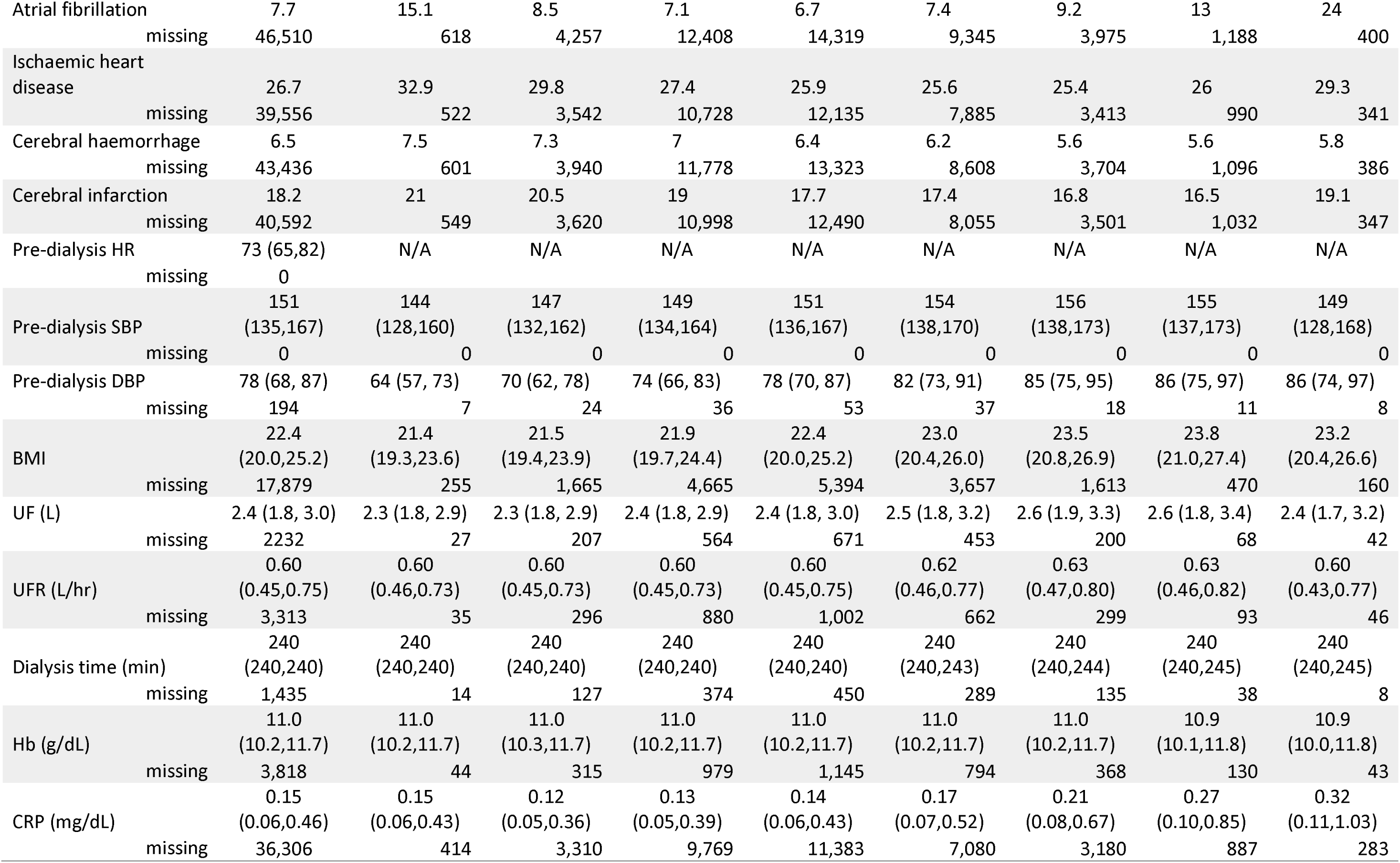
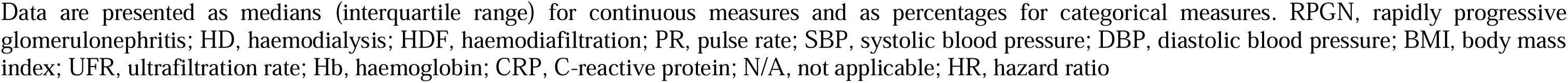
Baseline characteristics of the overall patients and those categorised by the pulse rate.

### Association of combined pre-HD SBP and PR with all-cause mortality

During 263,016 person-years of follow-up, 24,623 deaths were observed, with an incidence rate of 9.4 per 100 person-years. Descriptive statistics for all-cause mortality by the combined category of pre-HD SBP and PR are presented (Supplementary Table 1). The model for the combined SBP and PR category explained mortality better than the model that included the SBP and PR categories separately (*P* for interaction by the Wald test <0.001). The combined effects of SBP and PR on mortality in terms of HRs and RERI are shown in Figure 1. In subgroups with a PR of 60-<70 / min or lower, the increased HR of risk by SBP category was U-shaped (Figure 1A). Regardless of the PR subgroup, the association between low SBP and high all-cause mortality risk was consistent. A monotonic increase in HR with increasing PR was observed mainly in the subgroups with an SBP of 140-<160 mmHg or lower. In the subgroup with SBP >180 mmHg, the increased HR of risk by the PR category was U-shaped.

**Figure 1.**
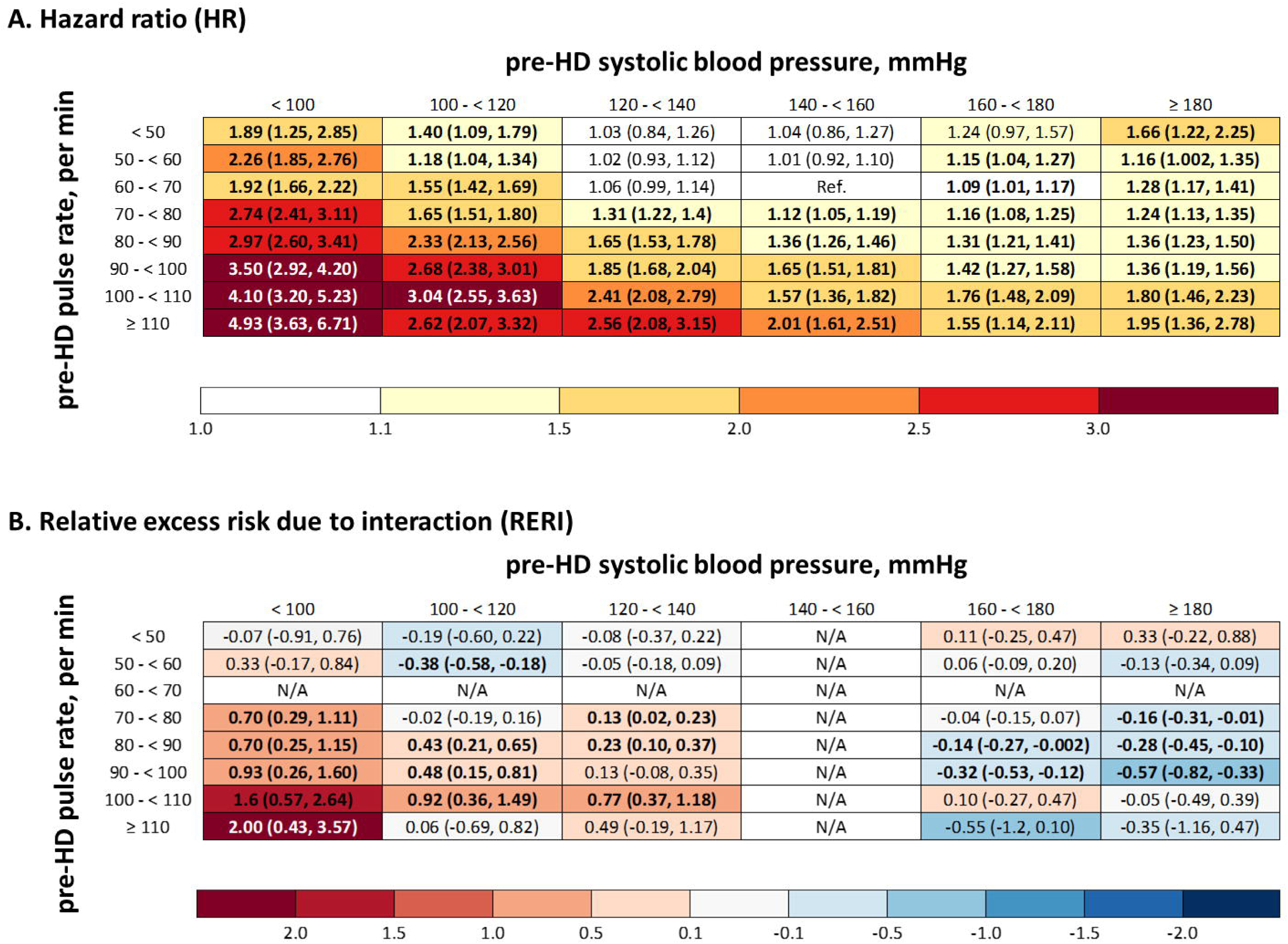
Heatmap showing the association between the combination of pre-haemodialysis systolic blood pressure and pulse rate and all-cause mortality. A. Hazard ratios; B. Relative excess risks due to interaction Hazard ratios and relative excess risks owing to interactions are estimated using the same Cox proportional hazards model. Pre-haemodialysis systolic blood pressure of 140-<160 mmHg and a pulse rate of 60-<70 / min is the reference category. The data are adjusted for age; sex; body mass index; serum C-reactive protein level; haemoglobin level; smoking status; use of antihypertensive drugs; history of atrial fibrillation, diabetes, ischaemic heart disease, cerebral infarction, and cerebral haemorrhage; interdialytic weight gain; and diastolic blood pressure. Each cell is coloured according to the numerical value of the point estimates. Numbers in parenthesis indicate 95% confidence intervals. HD, haemodialysis; N/A, not applicable

In addition, evidence of effect modification by additive scales was observed (Figure 1B). In particular, most categories with a lower pre-HD SBP or higher PR than the 120-<140 mmHg and 70-<80/min categories, respectively showed positive RERIs. For example, the strongest positive RERI was found in the SBP <100 mmHg and PR ≥110 / min category (RERI=2.00, 95% confidence interval [CI] 0.43-3.57). Conversely, many categories with a higher pre-HD SBP or PR than the 160-<180 mmHg and 70-<80/min categories showed negative RERIs.

### Association of combined pre-HD SBP and PR with cardiac mortality

During the same follow-up, 7,056 cardiac mortalities were observed with an incidence rate of 2.7 per 100 person-years. Descriptive statistics for cardiac mortality by the combined category of pre-HD SBP and PR are presented (Supplementary Table 2). The model for the combined SBP and PR categories explained cardiac mortality better than the model that included the SBP and PR categories separately (*P* for interaction by the Wald test: 0.002). The combined effects of SBP and PR on cardiac mortality in terms of HRs and RERI are shown in Figure 2. Similar to all-cause mortality, the increased HR by SBP category was U-shaped in subgroups with PRs of 60-<70/min or lower (Figure 2A). Regardless of the PR subgroup, the association between low SBP and high cardiac mortality risk was consistent. A monotonic increase in HR with increasing PR was mainly observed in subgroups with an SBP of 140-<160 mmHg or lower. In the subgroup with an SBP of 160-<180 mmHg or higher, the increased HR of risk by the PR category was U-shaped.

**Figure 2.**
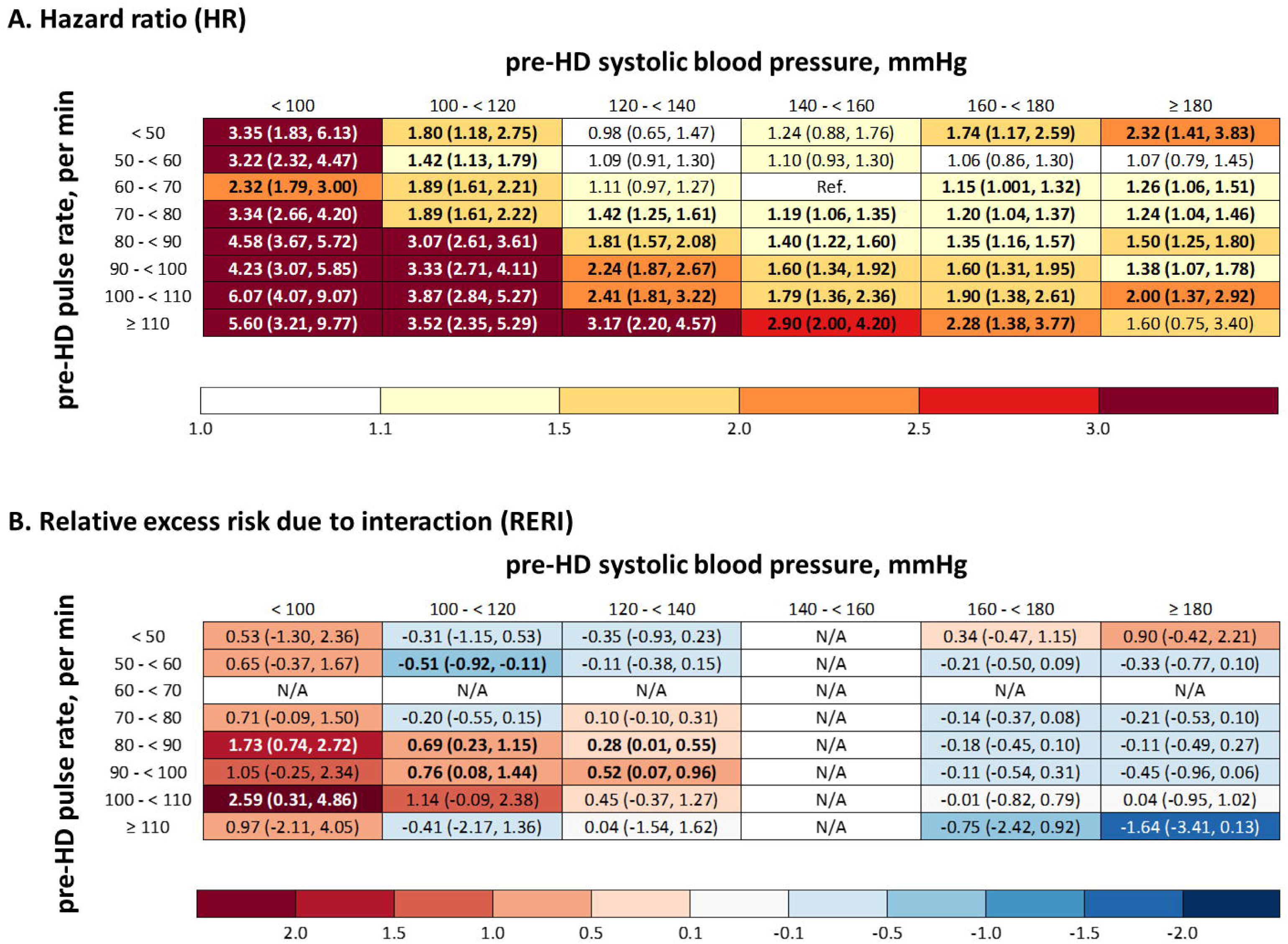
Heatmap showing the association between the combination of pre-haemodialysis systolic blood pressure and pulse rate and cardiac mortality. A. Hazard ratios; B. Relative excess risks due to interaction Hazard ratios and relative excess risks owing to interactions are estimated using the same Cox proportional hazards model. Pre-haemodialysis systolic blood pressure of 140-<160 mmHg and a pulse rate of 60-<70 / min is the reference category. The data are adjusted for age; sex; body mass index; serum C-reactive protein level; haemoglobin level; smoking status; use of antihypertensive drugs; history of atrial fibrillation, diabetes, ischaemic heart disease, cerebral infarction, and cerebral haemorrhage; interdialytic weight gain; and diastolic blood pressure. Each cell is coloured according to the numerical value of the point estimates. Numbers in parenthesis indicate 95% confidence intervals.

In addition, evidence of effect modification by additive scales was observed (Figure 2B). In particular, most categories with a lower pre-HD SBP or higher PR than the 120-<140 mmHg and 80-<90/min categories had positive RERIs. Conversely, many categories with a higher pre-HD SBP or PR than the 160-<180 mmHg and 70-<80/min categories showed negative RERIs, although they were not statistically significant.

## Discussion

Using data from a nationwide registry of Japanese patients undergoing maintenance HD, we found that the combined category of SBP and PR explained the one-year risk of all-cause and cardiovascular mortalities better than the separate categories of pre-dialysis SBP and PR. For the HR for all-cause mortality, compared with the reference range of pre-dialysis SBP and PR (140-160 mmHg and 60-70 bpm, respectively), either an increase or decrease in SBP alone (i.e. a U-shaped relationship) and an increase in PR alone were associated with an increased risk; however, the combination of lower SBP and higher PR demonstrated a greater risk than lower SBP or higher PR alone. This was also supported by the presence of many low SBP and high PR categories showing positive RERI. Similarly, for the HR for cardiovascular mortality, compared with the pre-dialysis reference range for SBP and PR, an increase or decrease in SBP alone (i.e. a U-shaped relationship) and an increase in PR alone were associated with an increased risk; however, the combination of low SBP and high PR showed a greater risk compared with low SBP or high PR alone. This was supported by the presence of several low SBP and high PR categories showing positive RERI.

The findings from our study partially concur with those of previous studies on the relationship between pre-dialysis SBP or PR and prognosis among patients undergoing HD and provide a new interpretation by combining the two indices. For all-cause and cardiovascular mortalities, a U-shaped increase in HR by SBP category was observed mainly in the subgroup with a PR of 60-<70/min or less. The SBP categories with particularly low risk in these subgroups ranged from 120 to 160 mmHg, consistent with the observed pre-dialysis SBP range of 130-160 mmHg associated with the lowest risk among the total participants in the previous cohort studies.^1,2,4,28^ The elevated pre-HD SBP may reflect both arterial stiffness and IDWG.^29^ Increased cardiac preload and afterload, through the combination of arterial stiffness and the Frank-Starling mechanism associated with elevated ventricular volume,^30^ increases myocardial oxygen demand, thereby precipitating the development of cardiovascular events.^29^ However, because we adjusted for weight gain and history of cardiovascular diseases, the increased risks associated with higher pre-HD SBP may reflect structural changes, such as endothelial dysfunction and hypertensive cardiomyopathy, which were not captured by the adjusted variables.

The observed relationships between low pre-HD SBP and high risks of all-cause and cardiovascular mortalities, regardless of the PR category, are supported by previous studies.^4,31^ The underlying pathophysiology of chronic hypotension among patients on dialysis may be well defined in some cases,^32^ but remains poorly understood.^33^

Cardiac disease with severely reduced left ventricular ejection fraction and valvular heart disease may also occur.^32^ Pathophysiological mechanisms such as autonomic dysregulation and excess activity of vasodilatory mediators have also been noted.^32,33^ For example, nitric oxide, a strong vasodilator, is increased in uraemia, and plasma levels of nitrites are positively correlated with years of dialysis.^32^ Long-term (usually 5 years) haemodialysis is common in chronic hypotension.^32^ While our study did not capture the abovementioned structural heart diseases, the excess activity of vasodilatory mediators may be involved in this population, as the group with an SBP <100 mmHg had a particularly long history of dialysis (median 7.0 years).

The monotonic increase in HR by an increasing PR category was observed mainly in the subgroup with an SBP of 140-<160 mmHg or less. The cause of tachycardia in patients undergoing HD may be multifactorial, including prodromal congestive heart failure and sympathetic nervous system activation.^6^ Indeed, the category with the highest PR of ≥110/min was characterised by baseline characteristics consistent with congestive heart failure: high prevalence of diabetes mellitus, AF, and IHD. However, this may reflect unmeasured tachyarrhythmic diseases or heart failure with low cardiac output because we adjusted for the characteristics mentioned above. In contrast, especially for cardiovascular mortality, a U-shaped increase in HR by PR category was observed mainly in the subgroup with an SBP of 160-<180 mmHg or higher. Bradycardia may reflect medications with negative chronotropic effects, cardiac disease with damage to the conduction system (e.g. narrowing of the right coronary artery), or electrolyte imbalance. However, because prescribing β-blockers to patients on HD may be associated with a favourable prognosis,^34^ underlying pathologies requiring its prescription may be the culprit for the poor prognosis associated with bradycardia.

A novel finding of this study was that the combined pre-HD SBP and PR category explained the risks of all-cause and cardiovascular mortalities better than the pre-HD SBP or PR categories alone. In particular, most categories with lower pre-HD SBP or higher PR than the 120-<140 mmHg and 70-<80/min categories showed positive RERIs, indicating that the combined pre-HD SBP and PR category accounted for more patients with excess risks than the single categories. In addition, this subset includes combined categories corresponding to an SI >0.7, which showed poor prognosis among patients with myocardial infarction,^12^ that is, those with an SBP <100 mmHg and 70-<80/min or higher PR, or those with an SBP of 100-<120 mmHg and 90-<100/min or higher PR. The underlying pathologies explaining these categories may include the aforementioned cardiac diseases with low cardiac output, excess activity of vasodilatory mediators, decreased extracellular fluid volume, and malnutrition.^6,32^ Indeed, the population with an SBP <100 mmHg was characterised by a relatively low BMI, high CRP level, and increased IHD, which is seen in the complex of protein-energy wasting, inflammation, and cardiovascular disease associated with unfavourable prognosis.^35^

In contrast, many categories with a higher pre-HD SBP or PR than the 160-<180 mmHg and 70-<80/min categories showed negative RERIs. These categories corresponded to high RPP categories; however, we were unable to verify the increased mortality risk associated with high RPP observed in non-dialysis CKD.^7^ However, the exact reason for this remains unclear. Although speculative, this may indicate the robustness of the cardiac system with functioning compensatory mechanisms; for example, a concomitant increase in SBP and a parallel increase in PR with increasing fluid volume.

Finally, positive, although not statistically significant, RERI in the ≥180 mmHg and <50 / min category indicates the presence of a high-risk subgroup that cannot be characterised by a high SI or RPP.

This study has several implications for dialysis providers and policymakers. First, the identification of subsets with high RERIs through the simultaneous assessment of pre-HD SBP and PR will help the limited healthcare staff within dialysis facilities to focus care and interventions on patients with an excess risk of cardiac or all-cause mortality.^36^ For example, targeted care can be designed, such as screening for cardiac diseases, investigating the causes of malnutrition and interventions, and adjusting dialysis regimens for haemodynamic stability. Second, the assessment using the combined categories of pre-HD SBP and PR offers the potential for stratification of high-risk patients that cannot be identified not only by the SBP or PR categories alone, but also by indices defined by RPP or SI alone. Specifically, patients with high SBP and high PR, low SBP and high PR, and high SBP and low PR were at higher risk than the reference category. In contrast, a high RPP (i.e. SBP × PR) can only risk stratify those with high SBP and PR, whereas a high SI (i.e. PR/SBP) can only risk stratify those with low SBP and high PR. In addition, the combination of SBP and PR is more convenient in clinical practice than calculating the RPP and SI. Indeed, the heatmaps successfully visualised the degree of risk in a semiquantitative manner.^25^ Therefore, our findings will serve as a useful reference for the preparation of blood pressure guidelines for patients on maintenance HD and will be a user-friendly resource for general dialysis practitioners.

This study had several limitations. First, as in international cohort studies, such as the Dialysis Outcomes and Practice Patterns Study (DOPPS), the SBP and PR measurements captured in the JSDT are not standardised. However, in the JSDT, SBP and PR are usually measured in the supine position while lying in bed during preparation for HD sessions.^6^ Second, several unmeasured factors, such as underlying structural heart disease, serum potassium level, and drugs with negative inotropic or chronotropic effects, may have influenced the relationship between pre-HD SBP, PR, and outcomes. Third, the follow-up period of 1 year in this study may not have been long enough to examine the impact of SBP and PR on mortality from atherosclerotic heart disease.

In conclusion, among patients on maintenance dialysis, the combined pre-HD SBP and PR categories were shown to identify excess risk groups for one-year all-cause and cardiac mortalities more effectively than either category alone. Longitudinal studies with longer follow-up periods are needed to examine the long-term effects of the combination of SBP and PR.

## Data Availability

Data will be available immediately after publication with no end dates. Data will be shared upon reasonable request to the corresponding author with permission from the JRDR investigators. Restrictions apply to the availability of the data analysed in this study to preserve patient confidentiality.

## Acknowledgements

We acknowledge the efforts of the members of the Subcommittee for JRDR Regional Cooperation and staff members of the dialysis facilities who participated in the survey and responded to the questionnaires.

## Prior presentation

Presented in part at the 69th Annual Meeting of the Japanese Society for Dialysis Therapy; 8 June 2024; Yokohama.

