## Supplementary Figure 1 for "Combined pre-dialysis systolic blood pressure and pulse rate assessment for all-cause and cardiovascular mortalities: A nationwide cohort study on patients undergoing haemodialysis"

**Supplementary Figure 1. Directed acyclic graphs**

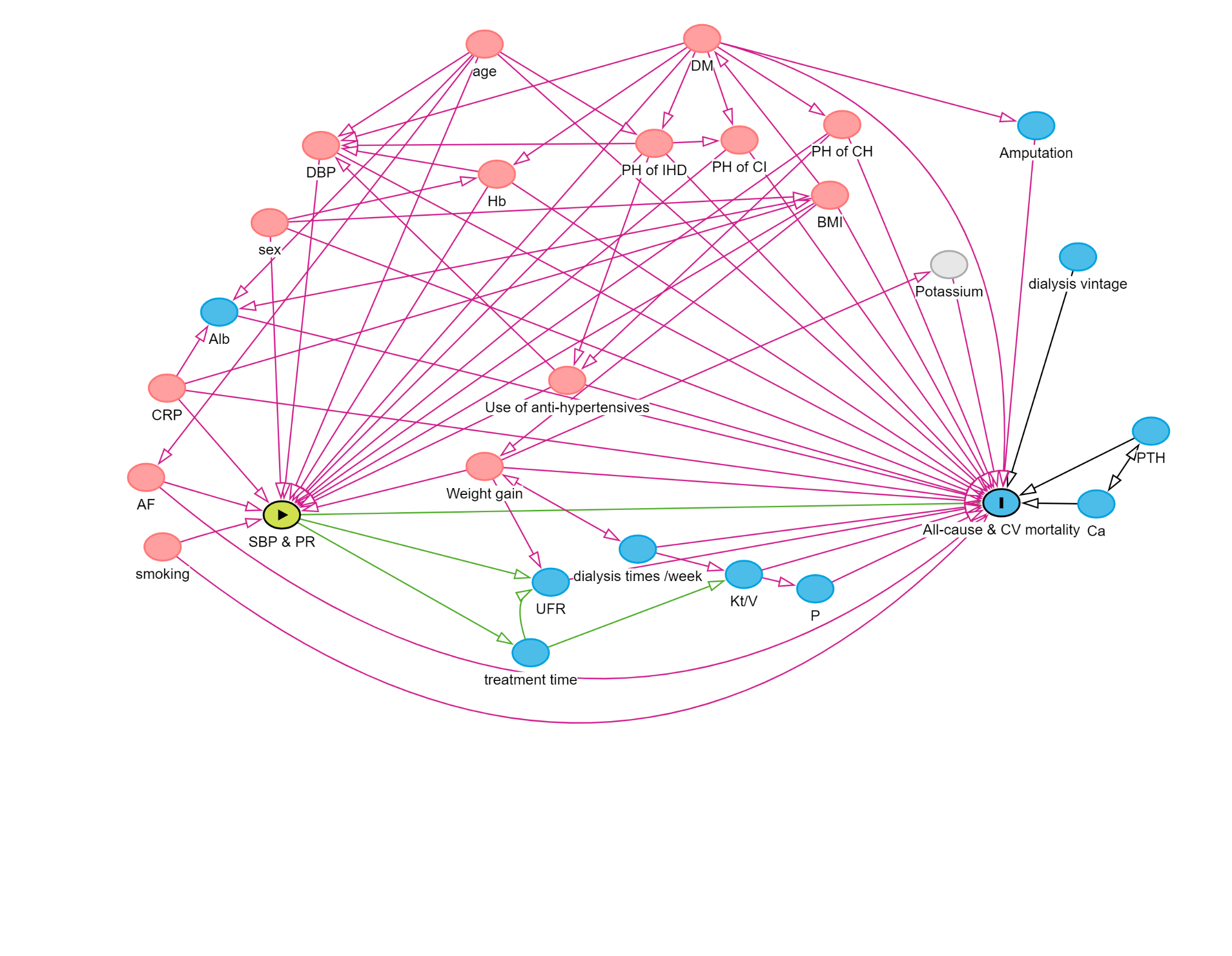

(A) The yellow-green oval indicates the combination of pre-HD SBP and PR ("exposure"), and the sky-blue oval indicates all-cause and cardiovascular mortalities ("outcomes"). The red circles represent the ancestors of both the exposure and outcome, whereas the light gray circles denote the unmeasured variables. The green line shows the causal pathway, starting with the exposure (combination of pre-HD SBP and PR), including only arrows moving away from the exposure, and ending with outcomes (all-cause and cardiovascular mortalities). The pink lines represent the biasing pathways. One of the minimally sufficient adjustment sets includes atrial fibrillation; body mass index; serum C-reactive protein levels; diastolic blood pressure; diabetes; haemoglobin level; use of antihypertensive drugs; interdialytic weight gain; age; sex; smoking; and history of ischaemic heart disease, cerebral haemorrhage, and cerebral infarction.

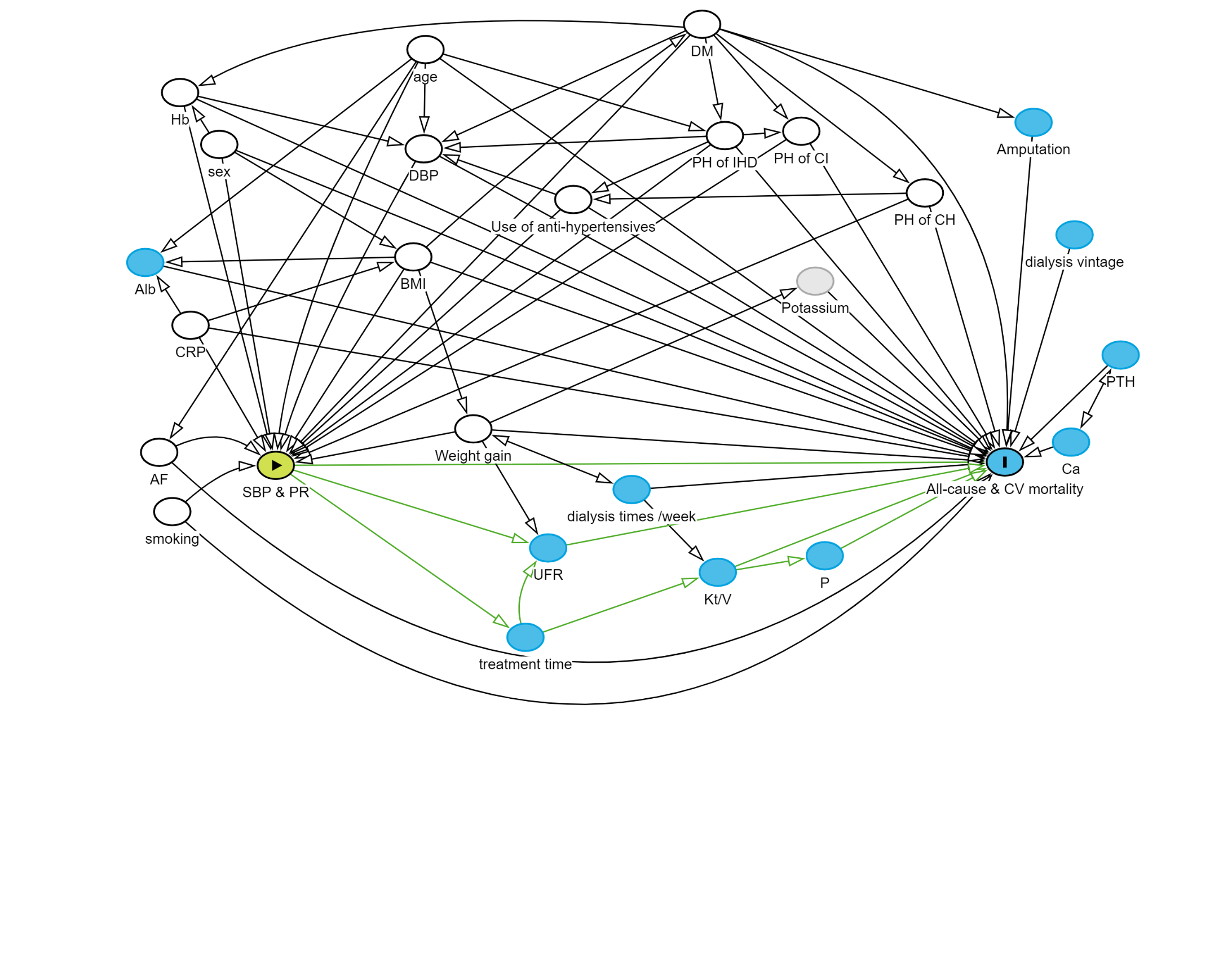

(B) With all variables in the minimally sufficient adjustment set adjusted ( white circles), only the green lines are displayed in the schema.

HD, haemodialysis; SBP, systolic blood pressure; PR, pulse rate
