## Supplementary Table 1 for "Combined pre-dialysis systolic blood pressure and pulse rate assessment for all-cause and cardiovascular mortalities: A nationwide cohort study on patients undergoing haemodialysis"

**Supplementary Table 1. Descriptive statistics on mortality by combined category of pre-dialysis systolic blood pressure and pulse rate**

|  |  | Systolic blood pressure category, mmHg | | | | | |
| --- | --- | --- | --- | --- | --- | --- | --- |
|  |  | <100 | 100-<120 | 120-<140 | 140-<160 | 160-<180 | ≥180 |
| Pulse rate category, per min | <50 | n = 101 | n = 399 | n = 919 | n = 1,108 | n = 609 | n = 283 |
|  |  | 24 (23.8%) | 68 (17%) | 97 (10.6%) | 107 (9.7%) | 69 (11.3%) | 42 (14.8%) |
|  |  | 90 pyr | 364 pyr | 869 pyr | 1055 pyr | 577 pyr | 265 pyr |
|  |  | 26.6 / 100 pyr | 18.7 / 100 pyr | 11.2 / 100 pyr | 10.1 / 100 pyr | 11.9 / 100 pyr | 15.9 / 100 pyr |
|  | 50-<60 | n = 431 | n = 2,329 | n = 6,759 | n = 8,605 | n = 5,174 | n = 2,098 |
|  |  | 110 (25.5%) | 279 (12%) | 600 (8.9%) | 697 (8.1%) | 471 (9.1%) | 193 (9.2%) |
|  |  | 370 pyr | 2,190 pyr | 6,468 pyr | 8,290 pyr | 4,959 pyr | 2,007 pyr |
|  |  | 29.7 / 100 pyr | 12.7 / 100 pyr | 9.3 / 100 pyr | 8.4 / 100 pyr | 9.5 / 100 pyr | 9.6 / 100 pyr |
|  | 60-<70 | n = 1,049 | n = 5,634 | n = 17,750 | n = 25,420 | n = 16,419 | n = 7,239 |
|  |  | 227 (21.6%) | 805 (14.3%) | 1,449 (8.2%) | 1,804 (7.1%) | 1,251 (7.6%) | 647 (8.9%) |
|  |  | 923 pyr | 5,216 pyr | 17,061 pyr | 24,583 pyr | 15,852 pyr | 6,940 pyr |
|  |  | 24.6 / 100 pyr | 15.4 / 100 pyr | 8.5 / 100 pyr | 7.3 / 100 pyr | 7.9 / 100 pyr | 9.3 / 100 pyr |
|  | 70-<80 | n = 1,197 | n = 5,514 | n = 18,178 | n = 28,937 | n = 20,701 | n = 10,318 |
|  |  | 297 (24.8%) | 778 (14.1%) | 1697 (9.3%) | 2018 (7%) | 1450 (7%) | 783 (7.6%) |
|  |  | 1,023 pyr | 5,099 pyr | 17,335 pyr | 28,001 pyr | 20,035 pyr | 9,965 pyr |
|  |  | 29.0 / 100 pyr | 15.3 / 100 pyr | 9.8 / 100 pyr | 7.2 / 100 pyr | 7.2 / 100 pyr | 7.9 / 100 pyr |
|  | 80-<90 | n = 836 | n = 3,498 | n = 10,401 | n = 17,550 | n = 14,601 | n = 8,210 |
|  |  | 263 (31.5%) | 684 (19.6%) | 1168 (11.2%) | 1366 (7.8%) | 1015 (7%) | 581 (7.1%) |
|  |  | 679 pyr | 3,101 pyr | 9,793 pyr | 16,855 pyr | 14,112 pyr | 7,933 pyr |
|  |  | 38.7 / 100 pyr | 22.1 / 100 pyr | 11.9 / 100 pyr | 8.1 / 100 pyr | 7.2 / 100 pyr | 7.3 / 100 pyr |
|  | 90-<100 | n = 408 | n = 1,585 | n = 4,277 | n = 6,853 | n = 6,269 | n = 4,221 |
|  |  | 133 (32.6%) | 362 (22.8%) | 543 (12.7%) | 622 (9.1%) | 431 (6.9%) | 265 (6.3%) |
|  |  | 322 pyr | 1,362 pyr | 3,984 pyr | 6,541 pyr | 6,068 pyr | 4,095 pyr |
|  |  | 41.3 / 100 pyr | 26.6 / 100 pyr | 13.6 / 100 pyr | 9.5 / 100 pyr | 7.1 / 100 pyr | 6.5 / 100 pyr |
|  | 100-<110 | n = 174 | n = 530 | n = 1,277 | n = 2,018 | n = 1,742 | n = 1,318 |
|  |  | 69 (39.7%) | 140 (26.4%) | 204 (16%) | 199 (9.9%) | 146 (8.4%) | 95 (7.2%) |
|  |  | 132 pyr | 438 pyr | 1,153 pyr | 1,906 pyr | 1,672 pyr | 1,266 pyr |
|  |  | 52.3 / 100 pyr | 32.0 / 100 pyr | 17.7 / 100 pyr | 10.4 / 100 pyr | 8.7 / 100 pyr | 7.5 / 100 pyr |
|  | ≥110 | n = 108 | n = 284 | n = 485 | n = 621 | n = 440 | n = 338 |
|  |  | 44 (40.7%) | 74 (26.1%) | 99 (20.4%) | 83 (13.4%) | 42 (9.5%) | 32 (9.5%) |
|  |  | 81 pyr | 243 pyr | 428 pyr | 577 pyr | 418 pyr | 321 pyr |
|  |  | 54.5 / 100 pyr | 30.5 / 100 pyr | 23.2 / 100 pyr | 14.4 / 100 pyr | 10.1 / 100 pyr | 10.0 / 100 pyr |

In each box surrounded by solid lines, the number of patients (n), risk and percentage, at-risk period (pyr, person-years), and incidence rate (/ pyr) are shown.
