## Supplementary Table 2 for "Combined pre-dialysis systolic blood pressure and pulse rate assessment for all-cause and cardiovascular mortalities: A nationwide cohort study on patients undergoing haemodialysis"

**Supplementary Table 2. Descriptive statistics on cardiac mortality by combined category of pre-dialysis systolic blood pressure and pulse rate**

|  |  | Systolic blood pressure category, mmHg | | | | | |
| --- | --- | --- | --- | --- | --- | --- | --- |
|  |  | <100 | 100-<120 | 120-<140 | 140-<160 | 160-<180 | ≥180 |
| Pulse rate category, per min | <50 | n = 101 | n = 399 | n = 919 | n = 1,108 | n = 609 | n = 283 |
|  |  | 11 (10.9%) | 23 (5.8%) | 25 (2.7%) | 34 (3.1%) | 26 (4.3%) | 16 (5.7%) |
|  |  | 90 pyr | 364 pyr | 869 pyr | 1,055 pyr | 577 pyr | 265 pyr |
|  |  | 12.2 / 100 pyr | 6.3 / 100 pyr | 2.9 / 100 pyr | 3.2 / 100 pyr | 4.5 / 100 pyr | 6.0 / 100 pyr |
|  | 50 -<60 | n = 431 | n = 2,329 | n = 6,759 | n = 8,605 | n = 5,174 | n = 2,098 |
|  |  | 41 (9.5%) | 90 (3.9%) | 169 (2.5%) | 201 (2.3%) | 113 (2.2%) | 46 (2.2%) |
|  |  | 370 pyr | 2,190 pyr | 6,468 pyr | 8,290 pyr | 4,959 pyr | 2,007 pyr |
|  |  | 11.1 / 100 pyr | 4.1 / 100 pyr | 2.6 / 100 pyr | 2.4 / 100 pyr | 2.3 / 100 pyr | 2.3 / 100 pyr |
|  | 60-<70 | n = 1,049 | n = 5,634 | n = 17,750 | n = 25,420 | n = 16,419 | n = 7,239 |
|  |  | 72 (6.9%) | 258 (4.6%) | 398 (2.2%) | 474 (1.9%) | 348 (2.1%) | 169 (2.3%) |
|  |  | 923 pyr | 5,216 pyr | 17,061 pyr | 24,583 pyr | 15,852 pyr | 6,940 pyr |
|  |  | 7.8 / 100 pyr | 4.9 / 100 pyr | 2.3 / 100 pyr | 1.9 / 100 pyr | 2.2 / 100 pyr | 2.4 / 100 pyr |
|  | 70-<80 | n = 1,197 | n = 5,514 | n = 18,178 | n = 28,937 | n = 20,701 | n = 10,318 |
|  |  | 96 (8%) | 235 (4.3%) | 479 (2.6%) | 566 (2%) | 396 (1.9%) | 210 (2%) |
|  |  | 1,023 pyr | 5,099 pyr | 17,335 pyr | 28,001 pyr | 20,035 pyr | 9,965 pyr |
|  |  | 9.4 / 100 pyr | 4.6 / 100 pyr | 2.8 / 100 pyr | 2.0 / 100 pyr | 2.0 / 100 pyr | 2.1 / 100 pyr |
|  | 80-<90 | n = 836 | n = 3,498 | n = 10,401 | n = 17,550 | n = 14,601 | n = 8,210 |
|  |  | 104 (12.4%) | 233 (6.7%) | 332 (3.2%) | 370 (2.1%) | 282 (1.9%) | 175 (2.1%) |
|  |  | 679 pyr | 3,101 pyr | 9,793 pyr | 16,855 pyr | 14,112 pyr | 7,933 pyr |
|  |  | 15.3 / 100 pyr | 7.5 / 100 pyr | 3.4 / 100 pyr | 2.2 / 100 pyr | 2.0 / 100 pyr | 2.2 / 100 pyr |
|  | 90-<100 | n = 408 | n = 1,585 | n = 4,277 | n = 6,853 | n = 6,269 | n = 4,221 |
|  |  | 42 (10.3%) | 115 (7.3%) | 169 (4%) | 158 (2.3%) | 131 (2.1%) | 74 (1.8%) |
|  |  | 322 pyr | 1,362 pyr | 3,984 pyr | 6,541 pyr | 6,068 pyr | 4,095 pyr |
|  |  | 13.0 / 100 pyr | 8.4 / 100 pyr | 4.2 / 100 pyr | 2.4 / 100 pyr | 2.2 / 100 pyr | 1.8 / 100 pyr |
|  | 100-<110 | n = 174 | n = 530 | n = 1,277 | n = 2,018 | n = 1,742 | n = 1,318 |
|  |  | 26 (14.9%) | 45 (8.5%) | 52 (4.1%) | 58 (2.9%) | 42 (2.4%) | 30 (2.3%) |
|  |  | 132 pyr | 438 pyr | 1,153 pyr | 1,906 pyr | 1,672 pyr | 1,266 pyr |
|  |  | 19.7 / 100 pyr | 10.3 / 100 pyr | 4.5 / 100 pyr | 3.0 / 100 pyr | 2.5 / 100 pyr | 2.4 / 100 pyr |
|  | ≥110 | n = 108 | n = 284 | n = 485 | n = 621 | n = 440 | n = 338 |
|  |  | 13 (12%) | 25 (8.8%) | 31 (6.4%) | 30 (4.8%) | 16 (3.6%) | 7 (2.1%) |
|  |  | 81 pyr | 243 pyr | 428 pyr | 577 pyr | 418 pyr | 321 pyr |
|  |  | 16.1 / 100 pyr | 10.3 / 100 pyr | 7.3 / 100 pyr | 5.2 / 100 pyr | 3.8 / 100 pyr | 2.2 / 100 pyr |

In each box surrounded by solid lines, the number of patients (n), risk and percentage, at-risk period (pyr, person-years), and incidence rate (/ pyr) are shown.
